# Prevalence and determinants of respiratory symptoms and functional disorders among children exposed to particulate matter through domestic and maternal occupational solid fuel use in Abidjan, Côte d’Ivoire – a cross-sectional study

**DOI:** 10.64898/2026.07.01.26357005

**Authors:** Auriane Pajot, Sonia Adjoua Dje, Flore Dick Amon Tanoh, Cathy Liousse, Thomas Thivillon, Madina Doumbia, Sylvain Gnamien, Yapo Marie, Michael Fayon, Véronique Yoboue, Olivier Marcy

## Abstract

**Background:** Children from low– and middle-income countries are particularly vulnerable to air pollution, a major environmental health risk, due to the immaturity of their lungs and their proximity to sources of household pollution. This study aimed to investigated the effect of exposure to biomass combustion through domestic and maternal occupational activities on respiratory health of children living in disadvantaged urban areas of Abidjan, Côte d’Ivoire.

**Methods:** Between February and December 2023, we conducted a cross-sectional observational study among children <16 years from households of women using biomass fuel for cooking (Group (G) 1), engaged in occupational fish smoking activities (G2), or primarily using gas for domestic cooking (G3). We assessed reported respiratory symptoms through standardized questionnaires and the presence of lung function impairments (LFI) though pulmonary function tests (spirometry and Rint). We assessed the association between study groups and key covariates with respiratory symptoms and LFI using mixed-effects regression models.

**Results:** Of 210 children enrolled – 119 (56.8%) female, median age 9 (6–12) years, 82 (39.0%) in G1, 47 (22.4%) in G2, and 81 (38.6%) in G3 – 15 (7.1%) reported wheezing in the last 12 months, 82 (39.0%) reported dry cough at night, 9 (4.9%) presented with dyspnea and 5 (2.7%) had chest pain on clinical examination, for an overall proportion of children with reported respiratory symptoms of 43.8% (92/210). Of 176 children who underwent pulmonary function testing, 59 (33.5%) had LFI detected, including 34 (45.9%) in G1, 8 (22.2%) in G2, and 17 (25.8%) in G3 (*p* = 0.011). Study group was associated with respiratory symptoms (G1 vs G3; aOR 3.82, 95% CI 1.68-8.68; p < 0.001), as well as with LFI (*p* = 0.042). Girls were at greater risk of LFI than boys (aOR 2.69, 95% CI 1.24-5.80; *p* = 0.012). Children whose mothers used charcoal or wood as cooking fuel had higher odds of respiratory symptoms (OR 2.61, 95% CI 1.22–5.58; *p* = 0.013) but no association was found with LFI (*p* = 0.459) compared with unexposed children.

**Conclusion:** Respiratory symptoms and lung function impairments were highly prevalent among children living disadvantaged, especially when mothers cook with wood or charcoal. Targeted maternal awareness and broader interventions to reduce household air pollution in disadvantaged urban areas are urgently needed to protect long-term respiratory health.

**ARTICLE SUMMARY:**

- **What is already known on this topic** – Low– and middle-income countries are particularly affected by ambient air pollution, and children are especially vulnerable. In these settings, biomass fuels are still widely used and are implicated as a risk factor for acute lower respiratory infections in children.
- **What this study adds** – This study suggests that the respiratory health of children living in disadvantaged settings is impaired with high prevalence of non-reversible OVDs, potentially indicating an early trajectory toward COPD at younger ages, and that exposure to charcoal and wood combustion suggest detrimental effects on both respiratory symptoms and pulmonary function, particularly in girls. Children living in households of fish smoking women however did not seem to be at higher risks of negative respiratory outcomes, probably due to limited exposure time in the smoking sites.
- **How this study might affect research, practice or policy** – Targeted awareness interventions for mothers, aiming to reduce household air pollution, as well as broader actions in disadvantaged urban areas, are needed to protect the long-term respiratory health of these children. The transition from charcoal to clean fuels such as gas or electricity for domestic use could help protect the health of children living in disadvantaged settings.

## INTRODUCTION

Considered the second leading risk factor for death in 2021, air pollution is a major global environmental health threat, disproportionately impacting populations in low– and middle– income countries (LMIC)(1). Despite expanded air quality monitoring in many cities worldwide in recent years, the vast majority of the global population remains exposed to this risk factor, with fewer than 1% of urban areas in LMIC meeting the WHO guidelines for ambient air quality (2). While levels of ambient fine particulate matter of 2.5µm (PM_2.5_) are declining or stabilizing in several world regions, they continue to rise across Africa (3). In sub-Saharan Africa, rapid urbanization, unregulated traffic emissions, open waste burning and the widespread use of biomass fuels remain major drivers of elevated fine particulate matter (PM_2.5_ and PM_10)_ pollution (4–12).

Children are particularly at risk of health consequences from air pollution, due to several factors. Physiological and anatomical factors lead to higher particle deposition in their lungs during a critical period of development when their organs are not yet mature (13,14). Moreover, children have a higher respiratory rate than adults, which means they inhale more particles (13,14). Several studies have shown that increased exposure to particulate matter in ambient air is associated with a rise in emergency admissions for respiratory diseases, including asthma in children (15,16) and increases the risk of pneumonia (17–20). Women’s exposure to various pollutants directly influences that of their children, who spend a large part of their early years in close proximity to them (21).

During the past fifteen years, extensive documentation and monitoring efforts have been conducted in Asia regarding air pollution, in contrast to Africa, where research remains limited despite both regions severely affected (1,22). In Côte d’Ivoire, which had an urbanization rate of 53% and an urban population growth of 3.4% in 2023, studies have estimated that urban PM exposure exceeds WHO guideline thresholds by three to ten times (6,23–25). After road traffic, domestic fires represented the second largest source of pollution in Abidjan, the capital city, accounting for 40% of total PM_2.5_ emissions (26), with higher PM_2.5_ concentrations in households using solid fuels such as wood or charcoal (12,27–29). In addition to the use of biomass fuel for cooking, informal economic activities such as artisanal fish smoking, further contribute to sustained indoor and outdoor air pollution, due to smoking sites located in close proximity to the dwellings(4–8,30). This traditional method of food preservation has been practiced for generations in West Africa and remains essential for food security, nutrition and household income (31). Recently, alarming real time 24-hour average PM_2.5_ exposure levels above 200-250 µg/m³ have been reported for both housewives using charcoal or wood for domestic cooking and fish-smoking women (32). Women are exclusively responsible for this smoking activity, often accompanied by their young children (30,31,33,34), exposing this highly vulnerable population to health consequences of air pollution.

Few researches have examined the contribution of maternal occupational biomass exposure and household fuel use to children’s respiratory health in sub-Saharan Africa, highlighting the need to better understand these determinants in urban, disadvantaged contexts (24,35). We hypothesized that children exposed to high levels of PM_2.5_ through use of biomass fuels for either domestic cooking or maternal occupational activities would present a higher burden of respiratory symptoms and lung function impairment (LFI). Here, we specifically sought to assess the impact of exposure to biomass combustion on respiratory health among three groups of children from households engaged in cooking using charcoal, in fish smoking activities, or mostly using gas for cooking, living in disadvantaged urban areas of Abidjan.

## METHODS

### Study design and population

We conducted a cross-sectional observational study from February to December 2023, within a pediatric extension of the APIMAMA project (2022–2026, https://apimama.org), which investigates domestic air pollution from biomass combustion and its respiratory effects in women in Abidjan, Côte d’Ivoire. APIMAMA focused on three groups of women highly exposed to biomass smoke: women using charcoal for cooking at home, fish-smoking women, and women involved in charcoal production. Using the APIMAMA participant registry, APIMAMA Kids included children under 16 years of age of women enrolled in two APIMAMA exposure groups: group 1 (G1) included children whose mother or grand-mother use charcoal for household cooking, G2 included children and grand-children of women engaged in fish smoking activities using wood. In addition, G3 was added as a specific control for this pediatric component, and included a convenience sample of children whose mother primarily use butane gas for cooking. For feasibility reasons, we were unable to include the children of women involved in charcoal production. Informed consent was obtained from legal guardians, and children aged ≥ 7 years provided assent. All adult participants were required to understand and communicate in French.

### Study settings

All participating households were located in Yopougon, the most populous commune in Abidjan with underserved, high-density areas characterized by unregulated housing and limited infrastructure. Each participant was selected based on predominant types of household or occupational biomass use within a low-income urban district of Yopougon for G1 and G3, and across the wider area for G2. (36,37). Consenting and data collection took place in participants’ residential environments, as well as in rooms provided by local chiefs or health centers in the different study neighborhoods for administering questionnaires and conducting respiratory function tests.

### Study procedures

Children’s health status was assessed using i) a respiratory health questionnaire derived from the International Study of Asthma and Allergies in Childhood (ISAAC), administered via tablet using REDCap software, ii) clinical assessments including measurement of vital signs and clinical examination, and iii) pulmonary function tests. The assessments were done by a trained nurse (questionnaires and spirometry) and pediatric pulmonologists (clinical examination and Rint).

Spirometry tests were performed on children aged ≥ 5 years using a spirometer (Spirolab, MIR France) and conducted and interpreted according to the Global Lung Initiative (GLI) reference equations and in line with American Thoracic Society (ATS) technical standards, with adaptations appropriate for pediatric populations (38,39). Measurements included forced vital capacity (FVC), forced expiratory volume in 1 second (FEV₁), forced mid-expiratory flow (FEF₂₅_-_₇₅) and the FEV₁/FVC ratio. Each child was required to perform at least two acceptable spirometry maneuvers (regular shape, free of artifacts, with an expiratory duration > 6 seconds, or > 3 seconds for children under 10 years, or a visible expiratory plateau ≥ 1 second, up to 8 attempts per child). The test was considered complete if the difference between the two best FVC and FEV1 values was < 150 mL (38). A bronchodilator reversibility test was also performed using salbutamol spray, 100 µg per puff. Spirometry data were extracted using the Winspiro PRO software at the end of the collection period. For children under 5 years or those unable to perform valid spirometry, airway resistance was assessed using the Airway Interruption Technique (Rint), a noninvasive and age-appropriate method, measured with a Spirodyn pneumotachograph, including a bronchodilator reversibility test.

Quantitative data on household characteristics, maternal occupation, cooking practices, housing features, and potential sources of air pollution were collected by trained field interviewers, using questionnaires, as part of the APIMAMA main study.

### Outcomes and variables of interest

Primary outcomes for this study were lung function impairment (LFI) and respiratory symptoms. LFI was defined as the presence of either obstructive (OVD) whether reversible or not, restrictive (RVD) or mixed ventilatory disorders (MVD) based on spirometry and Rint measurements. Non-reversible OVD was defined by a percent predicted forced mid-expiratory flow (ppFEF₂₅_-_₇₅) < 70% and/or ppFEV1 < 80%, with no reversibility after bronchodilator testing. RVD was defined by ppFVC < 80% and ppFEF₂₅_-_₇₅ and ppFEV1 ≥ 80%. MVD were characterized by FVC < 80% combined with pp FEF₂₅_-_₇₅ < 70% and/or ppFEV1 < 80%. Presumptive asthma was defined as a reversible OVD with a ppFEV1 or ppFVC improvement ≥ 12% after bronchodilatation. OVD as measured by Rint was defined as ppRint > 150% with reversibility of obstruction indicated by a ≥ 35% after bronchodilator testing. The presence of respiratory symptoms was defined as reporting the presence of at least one among: dyspnoea, chest pain, nocturnal cough, or wheezing within the past 12 months. The secondary outcomes were presumptive asthma on respiratory function testing, as described above, and as defined according to the ISAAC criteria, as a history of wheezing at any time in life with at least one episode of wheezing in the past 12 months, or a parental report of physician-diagnosed asthma.

The main independent variable of interest was the study group; other variables of interest were the age, sex, body mass index for age z-score, household income, use of charcoal for domestic cooking, and housing and living environment characteristics, such as the presence of at least one window in the house and the presence of animals indoors and the use of incense at home. The variable “smoking status” was not included in the analysis, as none of the participants reported being smokers.

### Data analyses

Based on low estimates of 2-3 children per mother/household enrolled in APIMAMA, the anticipated total sample size was 180–210 children across the three groups.

We described sociodemographic characteristics, housing conditions, sources of household air pollution, medical history, and respiratory function indicators using spirometric parameters and compared them across the 3 study groups using Student’s t-test or Kruskal-Wallis test, Pearson’s chi-squared test, Fisher’s exact test as appropriate.

We assess the association between study groups and other key covariates and 1) respiratory symptoms and 2) LFI using mixed-effect logistic models (glmer), taking into account the dependency among children from the same family through a random effect on the household identifier. The study group, sex and age were forced in the model. Variables of interest with a p-value < 0.25 in univariate analyses were included in multivariate models; and the final models were obtained using backward stepwise selection at a *p* = 0.05. Associations are reported as adjusted odds ratios (aOR) with 95% confidence intervals (95% CI). As cooking practices included mixing charcoal and gas and aiming to further understand the impact of biomass combustion beyond study group, we build similar models using any use of charcoal and/or wood as main explanatory variable, and including other covariates. Analyses were conducted using R, with statistical significance set at *p* < 0.05. Model assumptions were evaluated by inspecting diagnostic residual plots and conducting statistical tests for normality, homoscedasticity, and autocorrelation.

### Patient and public involvement

Children and their mothers contributed to optimizing sensor and spirometry placement and provided feedback on equipment acceptability. They also reported any issues during data collection, allowing prompt intervention. Results will be communicated in a format accessible to both children and adults.

## RESULTS

### Sociodemographic characteristics of children and sources of exposures

We enrolled 210 children from 87 families, of median age 9 (6–12) years, 119 (56.8%) of whom were female, and comprising 82 (39.0%) children from G1-charcoal/wood, 47 (11.4%) from G2-fish smoking and 81 (38.6%) from G3-gas. The proportion of female participants varied across groups with 52 (63.4%) girls in G1, 30 (63.8%) in G2 and 37 (45.7%) in G3 (p = 0.039) (Table 1 and S1 Appendix).

**Table 1.** – Demographics, medical characteristics and respiratory symptoms of study participants (N = 210)

| Characteristics | Overall<br>N = 210 | G1<br>Charcoal/wood<br>N = 82 | G2<br>Smoking fish<br>N = 47 | G3<br>Gas<br>N = 81 | P<br>value |
| --- | --- | --- | --- | --- | --- |
| Age (years) | 9 (6-12) | 9 (6-13) | 10 (5-12) | 8 (5-11) | 0.310 |
| Gender female | 119 (56.8) | 52 (63.4) | 30 (63.8) | 37 (45.7) | 0.039 |
| Educational level of the child |  |  |  |  | 0.628 |
| Young child, not at school | 42 (20.1) | 13 (15.8) | 8 (17.0) | 21 (26.2) |  |
| Preschool / Primary school | 112 (53.6) | 45 (54.9) | 25 (53.2) | 42 (52.5) |  |
| Middle school / High school | 46 (22.0) | 19 (23.2) | 12 (25.5) | 15 (18.7) |  |
| Employed | 5 (2.4) | 2 (2.4) | 2 (4.3) | 1 (1.2) |  |
| Unemployed/disabled | 4 (1.9) | 3 (3.7) | 0 | 1 (1.2) |  |
|  | N=183 | N=78 | N=37 | N=68 |  |
| Weight (kg) | 27 (19-41) | 27 (19-42) | 26 (19-39) | 27 (19-38) | 0.898 |
| Height (cm) | 130 (114-149) | 132 (116-151) | 130 (113-149) | 126 (111-146) | 0.460 |
| BMI | 16.3 (14.7-18.4) | 16.2 (14.6-17.6) | 16.4 (15.0-18.0) | 16.4 (14.6-19.4) | 0.696 |
| Dyspnea | 9 (4.9) | 4 (5.1) | 1 (2.7) | 4 (5.9) | 0.908 |
| Chest pain | 5 (2.7) | 4 (5.1) | 0 | 1 (1.5) | 0.280 |
| Resting vital signs | N= 138 | N=68 | N=25 | N=45 |  |
| SpO<sub>2</sub> (%) | 97 (95-98) | 97 (95-98) | 98 (96-99) | 97 (94-98) | 0.169 |
| Heart rate (beats/min) | 88 (76-98) | 86 (75-93) | 87 (76-100) | 93 (80-101) | 0.096 |
| Respiratory rate<br>(breaths/min) | 24 (22-28) | 27 (23-30) | 26 (24-28) | 22 (19-24) | <0.00<br>1 |
| Vital signs 1 minute post 3-minute step test | N=125 | N=67 | N=16 | N=42 |  |
| SpO<sub>2</sub> (%) | 96 (94-98) | 96 (94-98) | 98 (95-99) | 95 (92-98) | 0.090 |
| Heart rate (beats/min) | 99 (87-112) | 101 (86- 114) | 103 (87-110) | 95 (89-107) | 0.507 |
| Respiratory rate<br>(breaths/min) | 30 (27-34) | 33 (29-40) | 31 (28-35) | 27 (24- 30) | <0.00<br>1 |
|  | N = 210 | N = 82 | N = 47 | N = 81 |  |
| Noisy breathing/whistling in the<br>chest |  |  |  |  |  |
| Any time in the past | 45 (21.4) | 21 (25.6) | 10 (21.3) | 14 (17.3) | 0.432 |
| In the last 12 months | 15 (7.1) | 6 (7.3) | 4 (8.5) | 5 (6.2) | 0.941 |
| Asthma diagnosed | 20 (9.5) | 7 (8.5) | 6 (12.8) | 7 (8.6) | 0.691 |
| Dry cough at night (last 12<br>months) | 82 (39.0) | 49 (59.8) | 9 (19.1) | 24 (29.6) | <0.00<br>1 |
| Missed school due to<br>respiratory problem | 9 (4.3) | 5 (6.1) | 2 (4.3) | 2 (2.5) | 0.567 |
| Respiratory symptoms<sup>a</sup> | 92 (43.8) | 52 (63.4) | 11 (23.4) | 29 (35.8) | <0.00<br>1 |

Median (Q1-Q3); n (%)
Kruskal-Wallis rank sum test; Fisher's exact test; Pearson's Chi-squared test
(G1) children and grand children of women who use charcoal for cooking, (G2) children and grand children of women engaged in fish smoking activities, and (G3) children and grand children of women who use butane gas for cooking
<sup>a</sup>Respiratory symptoms defined as reporting at least one among: dyspnea, chest pain, nocturnal cough, or wheezing within the past 12 months

Among the 87 households with children enrolled, 71 (81.6%) had one window or more, which was frequent G1 (25/37, 67.6%) as compared with G2 (16/17, 94.1%) and G3 (30/33, 90.9%) (*p* = 0.018). Daily household income distribution differed significantly across groups (*p* < 0.001). Twenty-one (56.8%) households in G1 reported earning less than 3.54 USD (2000 FCFA) per day, compared with none in G2 and only two (6.1%) in G3. The majority of households in G3 (31, 93.9%) reported a daily income between 3.54 and 8.86 USD, whereas income above 8.86 USD/day was uncommon, observed mainly in G2 (4, 23.5%). Patterns of domestic cooking fuel use likewise differed across groups (*p* < 0.001). When considering any use of domestic cooking solid fuels (charcoal or wood), all women in G1 (37, 100%) reported exposure, compared with 12 (70.6%) in G2 and 4 (12.1%) in G3 (*p* < 0.001). Plastic waste was used for fire ignition by 12 households (22.6%) among those cooking with solid fuels, while 46 (86.8%) used pieces of dried natural rubber with a significant difference observed between groups (<0.001) (Table 2 and S2 Appendix).

**Table 2.** – Characteristics of household dwellings (N=87)

| Characteristic | Overall<br>N = 87 | G1<br>Charcoal/wood<br>N = 37 | G2<br>Smoking fish<br>N = 17 | G3<br>Gas<br>N = 33 | P value |
| --- | --- | --- | --- | --- | --- |
| Presence of at least 1 window in the house | 71 (81.6) | 25 (67.6) | 16 (94.1) | 30 (90.9) | 0.018 |
| Income (USD/day) |  |  |  |  | <0.001 |
| < 3,54 | 23 (26.4) | 21 (56.8) | 0 | 2 (6.1) |  |
| [3,54 – 8,86] | 57 (65.5) | 13 (35.1) | 13 (76.5) | 31 (93.9) |  |
| > 8,86 | 7 (8.0) | 3 (8.1) | 4 (23.5) | 0 |  |
| Kitchen in the house | 21 (24.1) | 7 (18.9) | 10 (58.8) | 4 (12.1) | 0.001 |
| Presence of animals at home | 20 (23.0) | 12 (32.4) | 1 (5.9) | 7 (21.2) | 0.097 |
| Type of fuel used for domestic cooking: |  |  |  |  |  |
| Charcoal | 53 (60.9) | 37 (100) | 12 (70.6) | 4 (12.1) | <0.001 |
| Wood | 5 (5.7) | 2 (5.4) | 2 (11.8) | 1 (3.0) | 0.418 |
| Gas | 65 (74.7) | 16 (43.2) | 16 (94.1) | 33 (100) | <0.001 |
| Charcoal or wood use yes/no | 53 (60.9) | 37 (100) | 12 (70.6) | 4 (12.1) | <0.001 |

Pearson's Chi-squared test; Fisher's exact test
(G1) children and grand children of women who use charcoal for cooking, (G2) children and grand children of women engaged in fish smoking activities, and (G3) children and grand children of women who use butane gas for cooking

### Children’s respiratory health

Noisy breathing or chest wheezing at any point in life was reported by 45 (21.4%) participants; 15 (7.1%) reported wheezing last 12 months and 9 (4.3%) reported missing school due to respiratory problems. Nocturnal dry cough last 12 months was reported in 82 (39.0%) children overall, with significant differences between groups: 49 (59.8%) in G1-charcoal/wood, 9 (19.1%) in G2-fish smoking, and 24 (29.6%) in G3-gas (p<0.001). Respiratory symptoms were reported by 92 participants (43.8%), with prevalence rates of 52 (63.4%) in Group G1, 11 (23.4%) in Group G2, and 29 (35.8%) in Group G3, respectively (p<0.001) (Table 1 and S1 Appendix).

Overall, 176 (83.8%) children underwent lung function testing, including 153 with spirometry and 23 with Rint. In children who performed spirometry, the median FVC and FEV₁ values were 1.63 (1.19–2.13) L and 1.58 (1.14–2.03) L, respectively. The median ppFVC and ppFEV₁ were significantly lower in G1 (81.8% and 88.2%, respectively), as compared with G2 (96.0% and 99.2%) and G3 (92.7% and 95.9%) (*p* < 0.001 and *p* = 0.002) (Table 3). Non-reversible OVDs were found in 22 (14.1%) children, without difference between groups (*p* = 0.111). Reversible OVD or asthma was found in 20 children (12.8%), without difference between groups (*p* = 0.712). RVDs were found in 15 children (9.8%), including 10 (15.6%), 0 and 5 (8.8%) in G1, G2, and G3, respectively (*p* = 0.034). Among the 23 children who underwent Rint measurements, 10 (43.5%) had abnormal values, with no significant differences between groups (*p* = 0.861). A reversible OVD was observed in 2 children (8.7%), including 1 in G1 (10.0%), and 1 in G3 (10.0%) (*p* = 0.999). Overall, LFI on pulmonary function test (N=176), were observed in 59 children (33.5%), with a higher proportion in G1 (34, 45.9%) compared with G2 (8, 22.2%) and G3 (17, 25.8%) (*p* = 0.011). Presumptive asthma was present in 22 children (12.5%), with no significant differences between groups (Table 3).

**Table 3.**
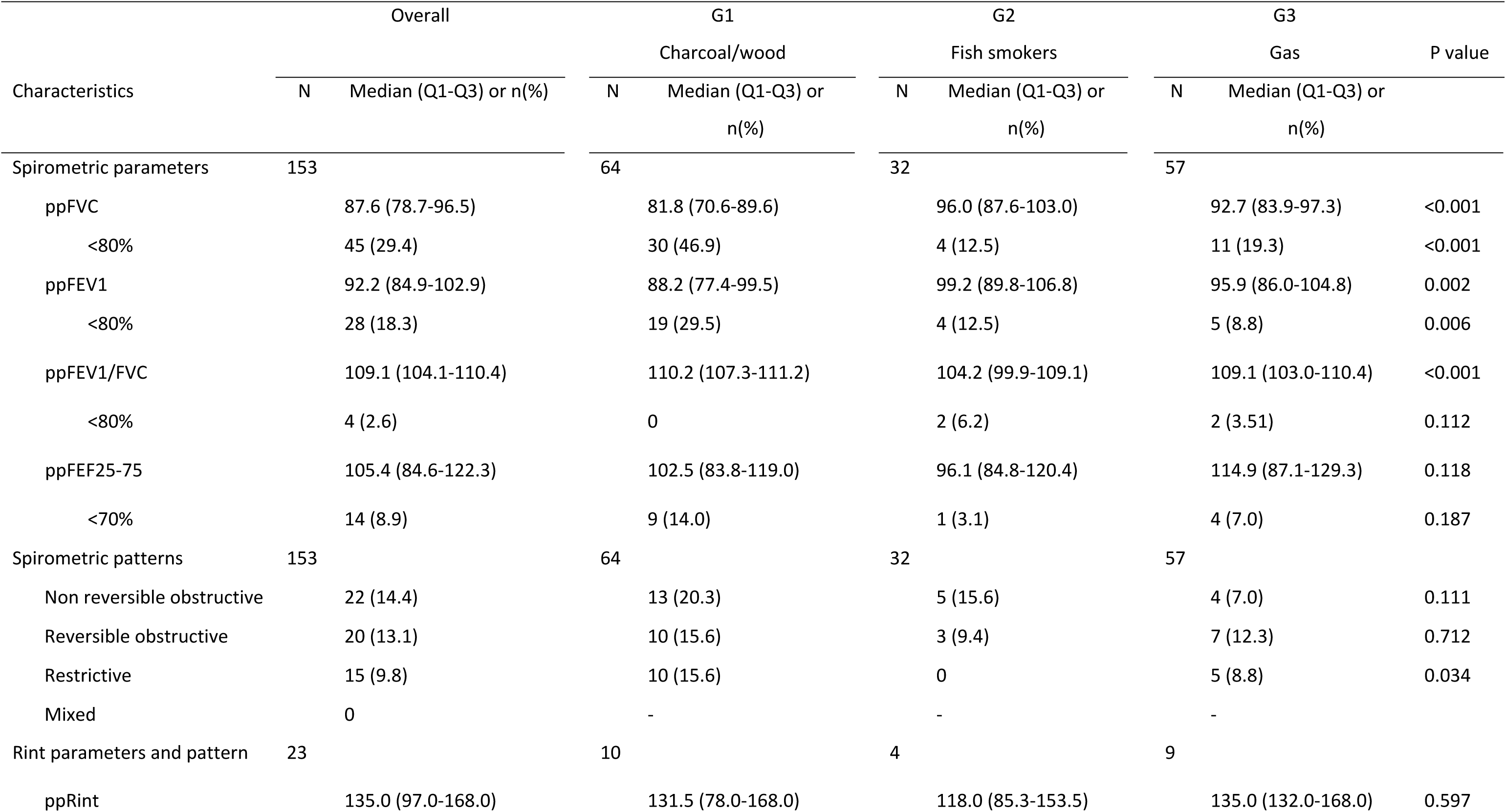
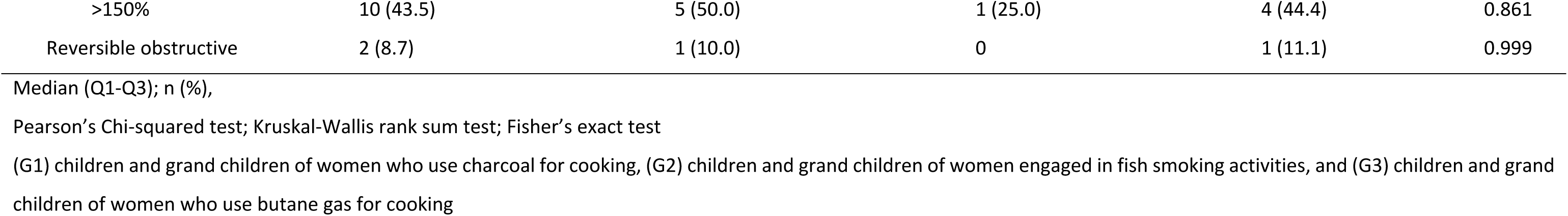
– Pulmonary function test and lung function impairment of children.

Respiratory symptoms were reported more frequently in children with LFI than in those without impairment (*p* = 0.041). Wheezing and nocturnal dry cough in the past 12 months were also more frequent in children with LFI (11.9% vs 3.4%, *p* = 0.044 and 54.2% vs 32.5%, *p* = 0.009, respectively). The association with wheezing was primarily observed in children with non-reversible OVDs (18.2% vs 4.6%, *p* = 0.038). Asthma reported was more commonly reported among those with non-reversible obstructive functional impairment (*p* = 0.047), but not among those with spirometry-based presumptive asthma. No other significant associations were observed between respiratory symptoms and lung function impairments (S3 Appendix).

### Association between exposure to biomass combustion and respiratory symptoms and LFI

In adjusted multivariate analyses, the risk of having respiratory symptoms was significantly associated with the study group (< 0.001). Children from G1 had more than three times the odds of presenting any respiratory symptoms as compared with children from G3 (aOR 3.82; 95% CI: 1.68–8.68; *p* < 0.001) (Table 4, S4 Appendix). G2 did not differ significantly from G3. There were no other variable significantly associated with respiratory symptoms. In a separate adjusted model excluding the study group, children living in households using wood or charcoal for cooking had more than twice the odds of respiratory symptoms compared with those using cleaner fuels (aOR 2.61, 95% CI 1.22–5.58; *p* = 0.013) (S4 and S5 Appendix).

**Table 4.**
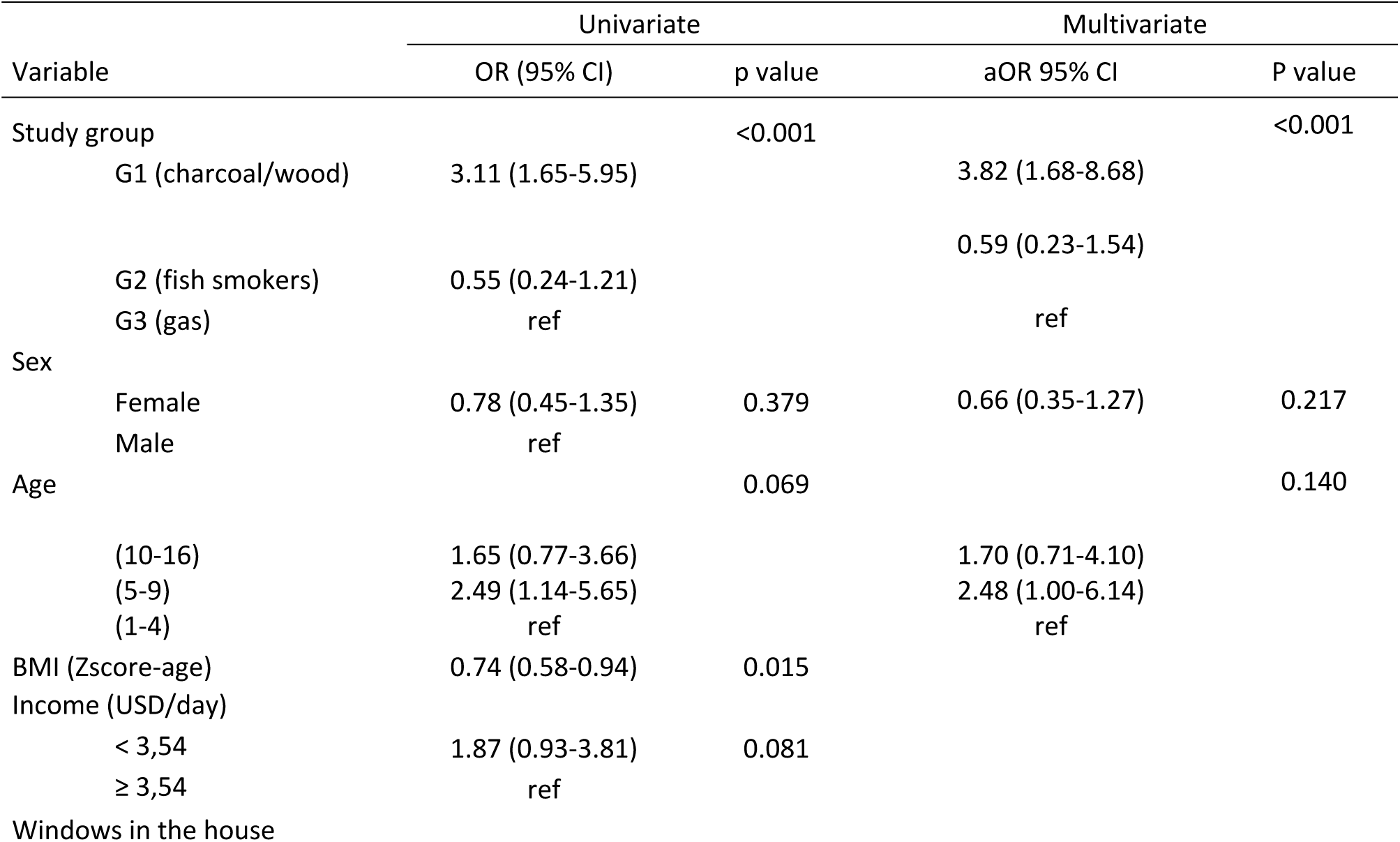
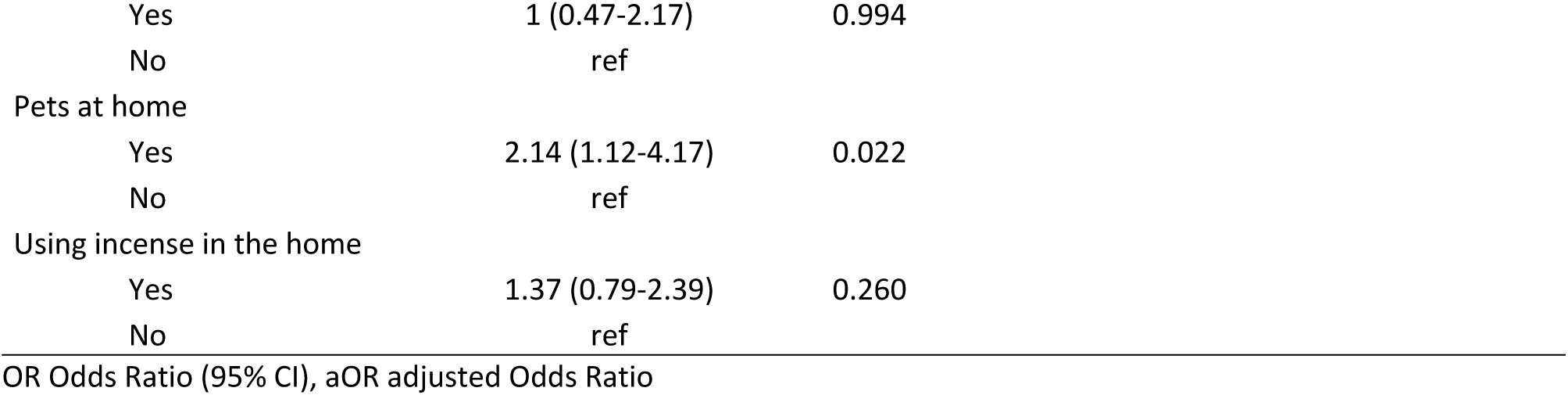
– Association between study group, key covariates and respiratory symptoms.

| Variable | Univariate |  | Multivariate |  |
| --- | --- | --- | --- | --- |
|  | OR (95% CI) | p value | aOR 95% CI | P value |
| Study group | | $<0.001$ | | $<0.001$ |
| G1 (charcoal/wood) | 3.11 (1.65-5.95) |  | 3.82 (1.68-8.68) |  |
| G2 (fish smokers) | 0.55 (0.24-1.21) |  | 0.59 (0.23-1.54) |  |
| G3 (gas) | ref |  | ref |  |
| Sex |  |  |  |  |
| Female | 0.78 (0.45-1.35) | 0.379 | 0.66 (0.35-1.27) | 0.217 |
| Male | ref |  |  |  |
| Age |  | 0.069 |  | 0.140 |
| (10-16) | 1.65 (0.77-3.66) |  | 1.70 (0.71-4.10) |  |
| (5-9) | 2.49 (1.14-5.65) |  | 2.48 (1.00-6.14) |  |
| (1-4) | ref |  | ref |  |
| BMI (Zscore-age) | 0.74 (0.58-0.94) | 0.015 |  |  |
| Income (USD/day) |  |  |  |  |
| < 3,54 | 1.87 (0.93-3.81) | 0.081 |  |  |
| ≥ 3,54 | ref |  |  |  |
| Windows in the house |  |  |  |  |

|  |  |  |
| --- | --- | --- |
| Yes | 1 (0.47-2.17) | 0.994 |
| No | ref |  |
| Pets at home |  |  |
| Yes | 2.14 (1.12-4.17) | 0.022 |
| No | ref |  |
| Using incense in the home |  |  |
| Yes | 1.37 (0.79-2.39) | 0.260 |
| No | ref |  |

OR Odds Ratio (95% CI), aOR adjusted Odds Ratio

In adjusted multivariate analyses, the G1-charcoal/wood group appears to be associated with an increased risk compared to G3-gas (aOR 2.11, 95% CI 0.92–4.86; p = 0.042) (Table 5). G2 did not differ significantly from G3. Female sex (aOR 2.69, 95% CI 1.24–5.80; p = 0.012), and older age (10–16 vs 1–9 years; aOR 2.83, 95% CI 1.36–5.90; p = 0.005) were also independently associated with LFI. A higher BMI-for-age-z-score showed a trend toward an inverse association with lung function impairment (p = 0.047) (Table 5 and S6 Appendix). In a separate adjusted model excluding the study group, living in households using wood or charcoal for cooking was not associated with increased odds of lung function impairment (aOR 1.34, 95% CI 0.61–2.94; p = 0.459). In contrast, a household income below USD 3.54 per day, being female, being aged 10–16 years, and a higher BMI-for-age z-score were associated with increased odds of lung function impairment (S7 Appendix).

**Table 5.**
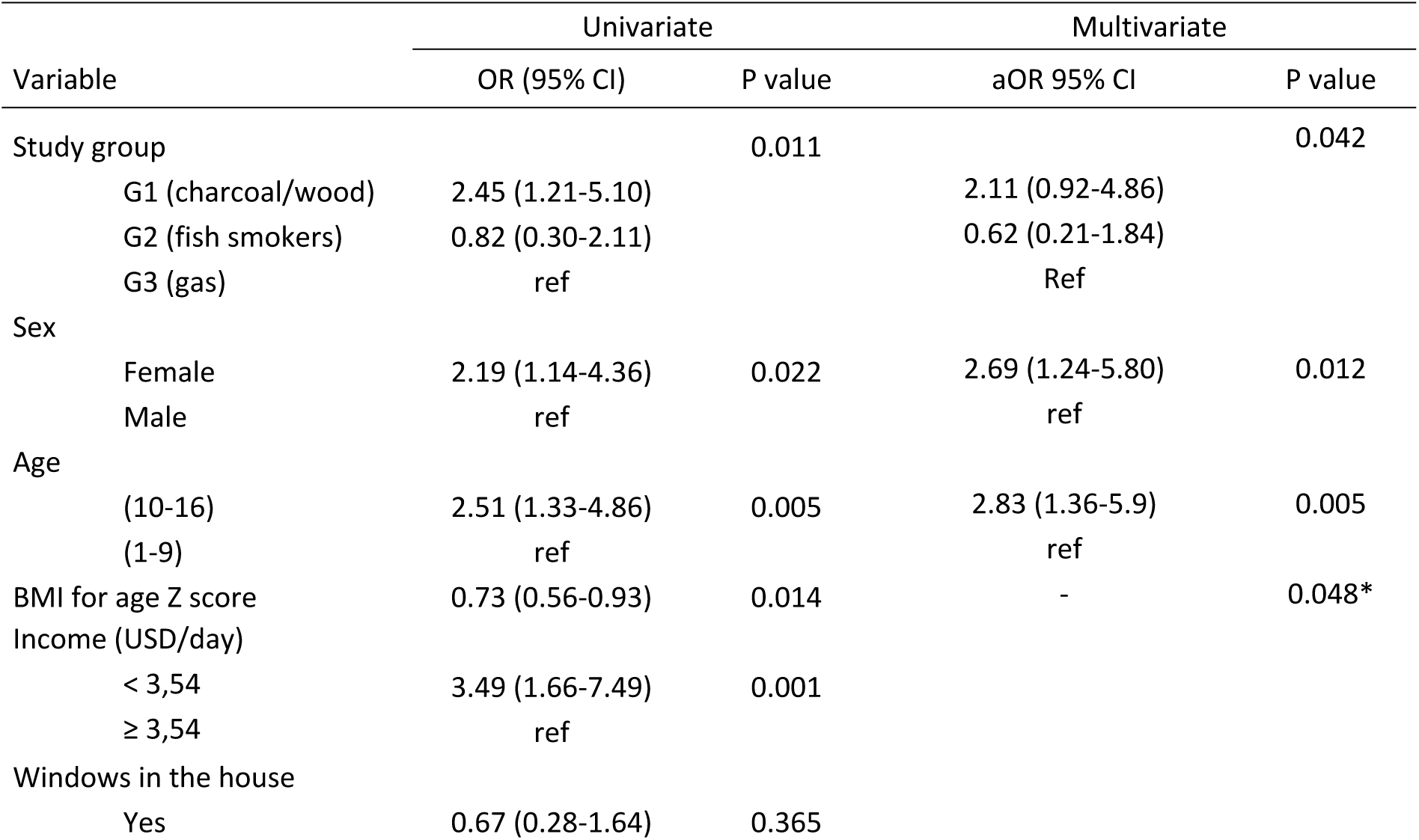
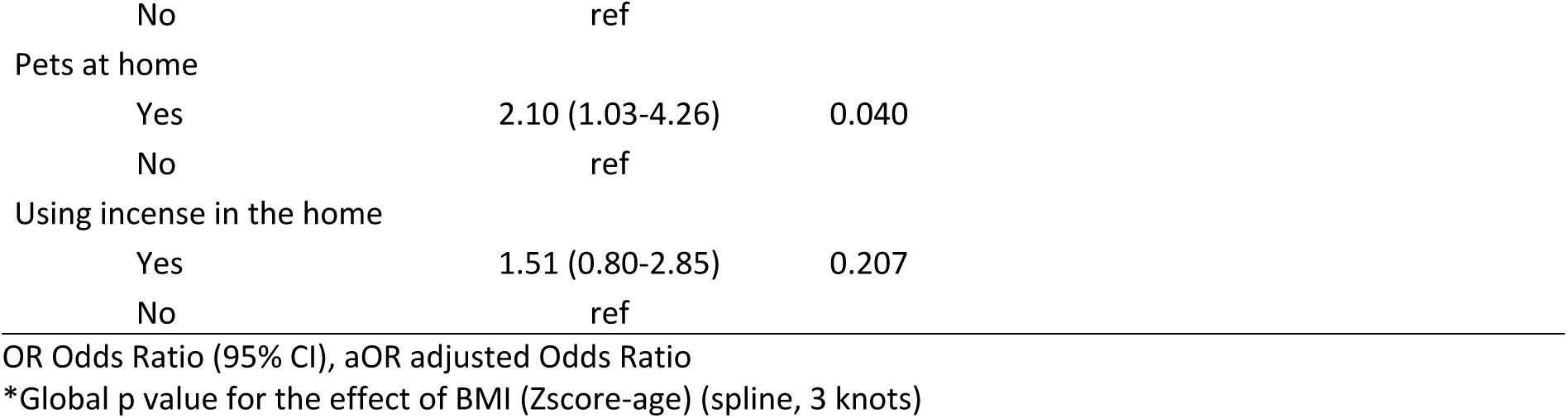
– Association between study group and key covariates and lung function impairments.

| Variable | Univariate |  | Multivariate |  |
| --- | --- | --- | --- | --- |
|  | OR (95% CI) | P value | aOR 95% CI | P value |
| Study group |  | 0.011 |  | 0.042 |
| G1 (charcoal/wood) | 2.45 (1.21-5.10) |  | 2.11 (0.92-4.86) |  |
| G2 (fish smokers) | 0.82 (0.30-2.11) |  | 0.62 (0.21-1.84) |  |
| G3 (gas) | ref |  | Ref |  |
| Sex |  |  |  |  |
| Female | 2.19 (1.14-4.36) | 0.022 | 2.69 (1.24-5.80) | 0.012 |
| Male | ref |  | ref |  |
| Age |  |  |  |  |
| (10-16) | 2.51 (1.33-4.86) | 0.005 | 2.83 (1.36-5.9) | 0.005 |
| (1-9) | ref |  | ref |  |
| BMI for age Z score | 0.73 (0.56-0.93) | 0.014 | - | 0.048* |
| Income (USD/day) |  |  |  |  |
| < 3,54 | 3.49 (1.66-7.49) | 0.001 |  |  |
| ≥ 3,54 | ref |  |  |  |
| Windows in the house |  |  |  |  |
| Yes | 0.67 (0.28-1.64) | 0.365 |  |  |

|  |  |  |
| --- | --- | --- |
| No | ref |  |
| Pets at home |  |  |
| Yes | 2.10 (1.03-4.26) | 0.040 |
| No | ref |  |
| Using incense in the home |  |  |
| Yes | 1.51 (0.80-2.85) | 0.207 |
| No | ref |  |

OR Odds Ratio (95% CI), aOR adjusted Odds Ratio
\*Global p value for the effect of BMI (Zscore-age) (spline, 3 knots)

## DISCUSSION

To our knowledge, this is among the first studies in West Africa to assess children living in precarious urban settings using both objective lung function measures and reported respiratory symptoms within a multi-source framework of pollution exposure. Our study showed a high burden of respiratory symptoms among children living in urban and disadvantaged areas. Pulmonary function testing, including spirometry and Rint measurements, detected impaired lung function in nearly one-third of those children, particularly those whose mothers used charcoal for household cooking (G1). Children in G1-wood/charcoal had a higher prevalence of respiratory symptoms and impaired lung function than those in the other groups. Maternal use of charcoal or wood was associated with respiratory symptoms. Girls exhibited a higher prevalence of functional respiratory disorders than boys.

The proportion of children reporting respiratory symptoms and those with lung function impairment was high in the study. Nocturnal dry cough, often considered a sensitive marker of bronchial irritation, was the most frequently reported symptom and was reported significantly more often among children with functional impairment, supporting the hypothesis of underlying bronchial inflammation or hyperresponsiveness (40). Wheezing in the previous 12 months, used to defined asthma by the ISAAC initiative was reported in less than one child out of 10. A meta-analysis estimated asthma prevalence at 13.9% (95% CI: 9.6–18.3) among children under 15 years in Africa (41) but estimates based on ISAAC questionnaires vary widely, from 5.6% in Djibouti (42), 12.7% in Madagascar (43) and 16.9-18.3% in Côte d’Ivoire (24,44). Such differences may reflect varying exposure levels, but also underreporting by mothers, linked to limited recognition of respiratory symptoms, stigmatization, or trivialization (45,46). Similar to other studies in LMICs, the prevalence of wheezing in our population exceeded the proportion of children previously diagnosed with asthma (S1 Appendix), suggesting substantial underdiagnosis in these settings (35,43). Contrary to expectations, symptoms of asthma reported through the ISAAC questionnaire were not declared more frequently among children with presumptive asthma identified on pulmonary function tests, possibly reflecting underreporting or underdiagnosis of asthma in some children, or a lack of symptom recognition by families. It should also be noted that asthma is characterized by reversible airflow limitation, consequently, the relationship between asthma and abnormal spirometry is not straightforward, as normal spirometric findings do not exclude the presence of asthma, as previously reported in the literature (47).

LFI detected in children including asthma, non-reversible obstructive and restrictive patterns, are consistent with emerging evidence on the effects of chronic exposure to biomass combustion. The predominance of obstructive patterns, particularly those with reversibility, points to active inflammation and bronchial hyperresponsiveness, likely driven by chronic irritation from, combustion-related toxic compounds and fine particulate matter (48–50). The reversibility of the observed impairment may be responsive to targeted public health interventions, in contrast to restrictive impairments, which are more likely to reflect structural or potentially irreversible alterations in lung function. Over time, persistent obstructive patterns in childhood may evolve into chronic expiratory flow limitation and increase the risk of chronic obstructive pulmonary disease in adulthood (51,52). Differences in LFI prevalence across studies likely reflect variations in age, exposure intensity, and socio-environmental conditions (49,50).

The study groups based on the main pattern of domestic or maternal occupational biomass combustion practices was associated with respiratory symptoms and LFI, highlighting different exposures to biomass combustion between groups of children enrolled in the study. As we had hypothesized, children enrolled in households using charcoal for cooking had higher risk of symptoms and LFI than children from households where gas is almost exclusively used. This is in line with previous large-scale studies in urban Bangladesh and Ghana that similarly reported that children from households relying on wood or charcoal were at higher risk of cough, wheezing and chest illnesses compared with those using cleaner fuels (53,54). Contrary to our initial hypothesis, children from households of women engaged in fish-smoking activities (G2) showed a low prevalence of adverse respiratory outcomes, comparable to that observed in G3, despite the more widespread use of biomass fuels for cooking in these households. This may reflect selection bias, as recruitment likely favored less-exposed, healthier children, and there may have been underreporting of symptoms due to the informal or even illegal nature of the women’s activities, depending on where they work (55). Households involved in fish-smoking also appeared to have relatively higher socioeconomic status, which could partly explain different levels of exposure of children to biomass combustion. As we showed elsewhere however female adolescents when helping their mothers are however exposed to very high levels of PM_2.5_ (56).

Our additional analyses accounting for fuel type rather than study group confirmed that exposure to wood and charcoal combustion compared to exclusive gas for domestic cooking, is independently associated with respiratory symptoms, highlighting the impact of household air pollution on children’s respiratory health. This finding is consistent with observations reported in the literature. Evidence from Nigeria further demonstrated that biomass fuel use increased the odds of severe asthma symptoms (OR = 2.37; 95% CI 1.16–4.84), reinforcing the role of domestic combustion as a determinant of respiratory morbidity in children (57). However, no association with LFI was observed for use of charcoal or wood after adjustment. Of note, this binary variable was solely defined on the basis of self-reported use of solid fuels for domestic cooking, without accounting for intensity or frequency of use. As a result, some participants may have been classified as exposed despite using charcoal or wood only marginally alongside gas. This exposure heterogeneity within the “exposed” group likely led to non-differential misclassification, which may have attenuated the true association between exposure and functional impairments and contributed to the null findings. It may also have failed to take into consideration the dose-response relationships that characterizes impairment versus symptoms that maybe more acute and linked to the irritative aspect of biomass combustion. Reporting bias may be present, as wood or charcoal users, aware of smoke-related risks, may over-report respiratory symptoms, while gas users may under-report them, perceiving their fuel as safer.

Beyond study groups and exposure to biomass combustion, other variables were independently associated with respiratory symptoms and LFIs. Girls were more likely than boys to present with functional respiratory disorders, even after adjusting for study group, age, and BMI for age Z-score. Although data from Africa are limited, a recent study in Thailand reported that among 93 children after adjusting for BMI, boys had significantly lower odds of any pulmonary abnormality compared with girls (adjusted OR = 0.084; 95% CI 0.017–0.417; p = 0.002) (50). In our context, this difference may be explained by the girls’ greater involvement in meal preparation and activities exposing them to smoke or roadside environments, which could increase their respiratory risk (21). These differences should also be examined while accounting for age and exposure to other pollutants, which may explain part of the observed variability (58). Low household income was associated with an increased risk of LFI. Although most households had minimal daily income (below 9 USD per day) those with poorer socio-economic conditions had a higher risk of LFIs, possibly due to higher in utero and life-long exposure to polluting practices, and more frequent acute respiratory infections. Likewise, a low BMI for age Z-score, reflecting undernutrition or malnutrition in children, which increases the risk of acute respiratory infections, heightened the likelihood of respiratory dysfunction compared with children with higher BMI. Socioeconomic status simultaneously shapes multiple dimensions of both environmental exposure and individual vulnerability, including the type of household fuel used, housing quality, and access to health resources. Low-income families are more likely to live in poorly ventilated dwellings, sometimes without windows, with visible dampness or mold, and in overcrowded conditions that further limit air circulation. In a subsample of children equipped with personal PM_2.5_ monitors, measured concentrations exceeded WHO recommendations by a wide margin (32). Low income therefore emerges as a central determinant, shaping environmental exposures, housing conditions, and nutritional status, and cumulatively contributing to impaired respiratory function in children (17,59).

Our study has several limitations. The absence of objective exposure data limits our ability to fully characterize dose-response relationships. Second, spirometry is a challenging exam in children, notably in younger ones and we used alternate definitions for OVDs. The small sample size of children who completed spirometry is also a limitation. This may have led to overestimating the prevalence of LFIs in the study. Third, symptoms were collected from mothers or children old enough to respond and were not clinically verifiable, which may introduce potential information bias. Future research should incorporate direct exposure measurements collected during the study, which will allow for a more precise characterization of the relationship between environmental exposures and respiratory outcomes. Although data were collected during the dry season, the timing within the season may affect lung function due to marked seasonal variability in ambient PM levels, temperature and humidity should be taken into account in future analyses (60).

Despite its limitations, the extensive data collected provided a detailed and representative description of living conditions in precarious neighborhoods in Abidjan, highlighting the high prevalence of respiratory symptoms and lung function impairments among children living in disadvantaged settings, particularly those whose mothers cook with charcoal at home. These findings highlight the need to implement clean cooking technologies to reduce household air pollution and improve respiratory health in vulnerable populations, with an initial priority being the promotion of clean household fuels, such as electricity and gas, to minimize domestic exposures. Our results underscore the importance of combining symptom assessment with functional test results, as well as the need for standardized international recommendations. Targeted awareness interventions for mothers to reduce household air pollution, together with broader public health actions in low-income urban areas, are needed to safeguard the long-term respiratory health of these children.

## ACKNOWLEDGEMENTS

The authors would like to thank the study participants, including the children and their families, for their valuable participation, as well as the students who contributed to data collection and analysis. They express their gratitude to the GPR IPORA (Interdisciplinary Policy-Oriented Research on Africa), the Francophone Respiratory Medicine Forum (EFP) and to AQuiRespi, a collaborative platform of the French Regional Health Agency (ARS) dedicated to respiratory health research and innovation, for their interest in the topic and support. They also thank the PAC-CI Research Center (ANRS site) in Abidjan for their logistical and technical support in implementing the study. Finally, we would like to thank the APIMAMA project, funded by the ANR, for its support in the implementation of this project.

## ETHICS STATEMENT

This study involves human participants. The study protocol was approved (N/Réf: 177-23/MSHPCMU/CNESVS-km) by the National Ethics Committee for Life Sciences and Health in Côte d’Ivoire (CNESVS). In addition, authorizations were obtained from local chiefs to conduct the study in each neighborhood. Participants gave informed consent to participate in the study before taking part.

## AUTHOR CONTRIBUTORS

AP wrote the study protocol, coordinated study implementation, contributed to conceptualization, data curation, methodology, and writing, collected data and drafted the first version of the manuscript. MY and SD were involved in data collection. CL, MD, FT, MF and VY contributed to conceptualization, methodology, supervision, and validation of the manuscript. SG and TT contributed to conceptualization and methodology. OM acted as guarantor, designed the study, supervised its scientific implementation and analysis, and contributed to manuscript write up and validation. All authors reviewed the manuscript and approved the final version.

## DATA AVAILABILITY

Data are available upon reasonable request. Data may be obtained from a third party and are not publicly available. All data relevant to the study are included in the article or uploaded as supplementary information. Data could be made available by study group to any researcher interested. Deidentified participant data and a data dictionary can be made available and shared under a data transfer agreement. Requests for access to the APIMAMA Kids study data should be sent to.

## FUNDING

This study received financial support from the French government through the University of Bordeaux’s France 2030 program / GPR IPORA (Interdisciplinary Policy-Oriented Research on Africa), https://ipora.africa/fr/. The project also benefited from a grant provided by the EFP (Établissement Francophone de Pneumologie). The funders had no role in the study design, data collection and analysis, decision to publish, or preparation of the manuscript.

## CONFLICTS OF INTEREST

Authors have declared that no competing interests exist.

## SUPPORTING INFORMATION

**S1 Appendix** – Demographics, medical characteristics and respiratory symptoms of study participants (N = 210)

**S2 Appendix** – Characteristics of household dwellings (N=87)

**S3 Appendix** – Association between reported respiratory symptoms and lung function impairment

**S4 Appendix** – Univariate analysis of the association between study group, demographic and housing characteristics on lung function impairment and respiratory symptoms

**S5 Appendix** – Association between the use of wood/charcoal for domestic cooking, key covariates and respiratory symptoms

**S6 Appendix** – Graph of predicted probability of lung function impairment by BMI-for-Age Z-Score

**S7 Appendix** – Association between the use of wood/charcoal for domestic cooking and key covariates and lung function impairments

## Notes

### Competing Interest Statement

The authors have declared no competing interest.

## REFERENCES

1. Health Effects Institute, Institute for Health Metrics and Evaluation’s Global Burden of Disease. State of Global Air Report 2024 | State of Global Air [Internet]. 2024 [cité 24 sept 2024]. Rapport No. Disponible sur: https://www.stateofglobalair.org/resources/report/state-global-air-report-2024

2. OMS. Des milliards de personnes respirent toujours un air pollué: nouvelles données de l’OMS [Internet]. 2022 [cité 8 avr 2024]. Disponible sur: https://www.who.int/fr/news/item/04-04-2022-billions-of-people-still-breathe-unhealthy-air-new-who-data

3. Brauer M, Freedman G, Frostad J, van Donkelaar A, Martin RV, Dentener F, et al. Ambient Air Pollution Exposure Estimation for the Global Burden of Disease 2013. Environ Sci Technol. 5 janv 2016;50(1):79–88. doi:10.1021/acs.est.5b03709

4. Keita S, Liousse C, Assamoi EM, Doumbia T, N’Datchoh ET, Gnamien S, et al. African anthropogenic emissions inventory for gases and particles from 1990 to 2015. Earth Syst Sci Data. 29 juill 2021;13(7):3691–705. doi:10.5194/essd-13-3691-2021

5. Doumbia E, Liousse C, Corinne GL, Ndiaye S, Diop B, Ouafo M, et al. Real time black carbon measurements in West and Central Africa urban sites. Atmos Environ. 1 juill 2012;54:529–37. doi:10.1016/j.atmosenv.2012.02.005

6. Gnamien S, Yoboue V, Liousse C, Ossohou M, Keita S, Bahino J, et al. Particulate Pollution in Korhogo and Abidjan (Cote d’Ivoire) during the Dry Season. Aerosol Air Qual Res. 1 janv 2020;20. doi:10.4209/aaqr.2020.05.0201

7. Brisoux L, Elgorriaga P. LES ENJEUX DE LA GESTION DES DECHETS A ABIDJAN La vitrine de la Côte d’Ivoire face aux défis de l’insalubrité [Internet]. 2018 [cité 9 mai 2022]. Rapport No. Disponible sur: https://documents.plateforme-re-sources.org/wp-content/uploads/2018/07/A283-Gestion-des-dechets-a-Abidjan-comp.pdf

8. Müller PE, Böni H. Les déchets solides municipaux en Afrique de l’Ouest: entre pratiques informelles, privatisation et amélioration du service public. 2012;7.

9. Kafando B, Windinpsidi Savadogo P, Sana A, Bagnoa V, Sanon S, Kouanda S, et al. Pollution intérieure par les PM2,5 issues des combustibles utilisés pour la cuisson des repas et risques sanitaires dans la ville de Ouagadougou. Environ Risques Santé. 2019;18(3):245–53.

10. Adon AJ, Liousse C, Doumbia ET, Baeza-Squiban A, Cachier H, Léon JF, et al. Physico-chemical characterization of urban aerosols from specific combustion sources in West Africa at Abidjan in Côte d’Ivoire and Cotonou in Benin in the frame of the DACCIWA program. Atmospheric Chem Phys. 6 mai 2020;20(9):5327–54. doi:10.5194/acp-20-5327-2020

11. Liousse C, Assamoi E, Criqui P, Granier C, Rosset R. Explosive growth in African combustion emissions from 2005 to 2030. Environ Res Lett. mars 2014;9(3):035003. doi:10.1088/1748-9326/9/3/035003

12. Lim S, Said B, Zurba L, Mosler G, Addo-Yobo E, Adeyeye OO, et al. Characterising sources of PM2·5 exposure for school children with asthma: a personal exposure study across six cities in sub-Saharan Africa. Lancet Child Adolesc Health. janv 2024;8(1):17–27. doi:10.1016/S2352-4642(23)00261-4 PubMed PMID: 38000380; PubMed Central PMCID: PMC10716619.

13. UNICEF. Les dangers de la pollution de l’air sur les enfants [Internet]. 2021 [cité 12 mars 2022]. Disponible sur: https://www.unicef.fr/dossier/pollution-de-lair

14. Sturm R. Theoretical models of carcinogenic particle deposition and clearance in children’s lungs. J Thorac Dis. 2012;4(4). doi:10.3978/j.issn.2072-1439.2012.08.03

15. Weichenthal SA, Lavigne E, Evans GJ, Godri Pollitt KJ, Burnett RT. Fine Particulate Matter and Emergency Room Visits for Respiratory Illness. Effect Modification by Oxidative Potential. Am J Respir Crit Care Med. sept 2016;194(5):577–86. doi:10.1164/rccm.201512-2434OC

16. Iskandar A, Andersen ZJ, Bønnelykke K, Ellermann T, Andersen KK, Bisgaard H. Coarse and fine particles but not ultrafine particles in urban air trigger hospital admission for asthma in children. Thorax. 1 mars 2012;67(3):252–7. doi:10.1136/thoraxjnl-2011-200324 PubMed PMID: 22156960.

17. WHO. Pollution de l’air à l’intérieur des habitations et santé [Internet]. 2022 [cité 18 janv 2022]. Disponible sur: https://www.who.int/fr/news-room/fact-sheets/detail/household-air-pollution-and-health

18. Smith KR, McCracken JP, Weber MW, Hubbard A, Jenny A, Thompson LM, et al. Effect of reduction in household air pollution on childhood pneumonia in Guatemala (RESPIRE): a randomised controlled trial. Lancet Lond Engl. 12 nov 2011;378(9804):1717–26. doi:10.1016/S0140-6736(11)60921-5 PubMed PMID: 22078686.

19. Kinney, lee, Asante. Prenatal and Postnatal Household Air Pollution Exposures and Pneumonia Risk: Evidence From the Ghana Randomized Air Pollution and Health Study. Chest. 1 nov 2021;160(5):1634–44. doi:10.1016/j.chest.2021.06.080

20. Lee AG, Kaali S, Quinn A, Delimini R, Burkart K, Opoku-Mensah J, et al. Prenatal Household Air Pollution Is Associated with Impaired Infant Lung Function with Sex-Specific Effects. Evidence from GRAPHS, a Cluster Randomized Cookstove Intervention Trial. Am J Respir Crit Care Med. 15 mars 2019;199(6):738–46. doi:10.1164/rccm.201804-0694OC PubMed PMID: 30256656; PubMed Central PMCID: PMC6423100.

21. WHO. Pollution de l’air et santé de l’enfant: prescrire un air sain: résumé [Internet]. Organisation mondiale de la Santé; 2018 [cité 18 janv 2022]. Rapport No.: WHO/CED/PHE/18.01. Disponible sur: https://apps.who.int/iris/handle/10665/275547

22. IQAir. 2024 World Air Quality Report | IQAir [Internet]. 2024 [cité 17 juin 2025]. Rapport No. Disponible sur: https://www.iqair.com/world-air-quality-report

23. Banque Mondiale. World Bank Open Data [Internet]. 2024 [cité 13 nov 2024]. World Bank Open Data. Disponible sur: https://data.worldbank.org

24. Pajot A, Yapo M, Coulibaly S, Doumbia M, Gnamien S, Kouao K, et al. Air pollution exposure, respiratory consequences, and perceptions among urban African children living in poor conditions – A case study in Abidjan, Côte d’Ivoire. PLOS Glob Public Health. 29 avr 2025;5(4):e0003703. doi:10.1371/journal.pgph.0003703

25. Gnamien S, Liousse C, Keita S, Silué S, Bahino J, Gardrat E, et al. Chemical characterization of urban aerosols in Abidjan and Korhogo (Côte d’Ivoire) from 2018 to 2020 and the identification of their potential emission sources. Environ Sci Atmospheres. 2023;3(12):1741–57. doi:10.1039/D3EA00131H

26. Gnamien S. Caractérisation de la pollution particulaire (PM10 et PM2.5) a Abidjan et Korhogo (Côte d’Ivoire) en lien avec la sante des populations [Internet]. 2022. Disponible sur: https://hal.science/tel-03811519v1/file/Th%C3%A8se%20de%20Gnamien%20N%27Douffou%20Konan%20Sylvain.pdf

27. Zhou Z, Dionisio KL, Verissimo TG, Kerr AS, Coull B, Howie S, et al. Chemical Characterization and Source Apportionment of Household Fine Particulate Matter in Rural, Peri-urban, and Urban West Africa. Environ Sci Technol. 21 janv 2014;48(2):1343–51. doi:10.1021/es404185m

28. Kouao AKR, Yoboue V, Silue S. Exposure to indoor and outdoor air pollution among children under five years old in urban area. 2020. doi:10.22034/gjesm.2019.02.00

29. Shupler M, Tawiah T, Nix E, Baame M, Lorenzetti F, Betang E, et al. Household concentrations and female and child exposures to air pollution in peri-urban sub-Saharan Africa: measurements from the CLEAN-Air(Africa) study. Lancet Planet Health. 1 févr 2024;8(2):e95–107. doi:10.1016/S2542-5196(23)00272-3 PubMed PMID: 38331535.

30. Monney UY, Diaby V, Bla BK, Konan ANKG, Yapo AF. Analyse socio-sanitaire du fumage de poisson dans la ville d’Abidjan (Côte d’Ivoire). Int J Biol Chem Sci. 2021;15(6):6. doi:10.4314/ijbcs.v15i6.8

31. Rivier, Kebe, Goli. Fumage de poissons en Afrique Ouest pour les marchés locaux et d’exportation [Internet]. 2009 [cité 24 mars 2022]. Rapport No. Disponible sur: https://www.doc-developpement-durable.org/file/Energie/biomasse-bois/fumoir/Fumage%20de%20poissons%20en%20Afrique%20Ouest_Rivier.pdf

32. Pajot A, Yapo M, Liousse C, Doumbia M, Gnamien S, Ahoua S, et al. Feasibility and results of joint ambulatory monitoring of exposure to particulate matter pollution and lung function in children in Abidjan, Côte d’Ivoire: a cross-sectional observational study. BMJ Open. 15 févr 2026;16(2):e109615. doi:10.1136/bmjopen-2025-109615 PubMed PMID: 41692517.

33. Dossou-Yovo P, Bokosssa I, Ahouandjinou H, Zolotokopova S, Alaguina I. Performance d’un dispositif amélioré de séchage de poisson fermenté appelé lanhouin au Bénin. Int J Biol Chem Sci. 2010;4(6):6. doi:10.4314/ijbcs.v4i6.64973

34. Djessouho DOC. Analyse socio-économique du fumage du poisson de la pêche artisanale maritime sur le littoral du Bénin. 2015;56.

35. Meme H, Amukoye E, Bowyer C, Chakaya J, Das D, Dobson R, et al. Asthma symptoms, spirometry and air pollution exposure in schoolchildren in an informal settlement and an affluent area of Nairobi, Kenya. Thorax. nov 2023;78(11):1118–25. doi:10.1136/thorax-2023-220057 PubMed PMID: 37280096; PubMed Central PMCID: PMC10715514.

36. CICG. ABIDJAN.DISTRICT.CI [Internet]. 2014 [cité 19 mars 2024]. Portail officiel du district d’abidjan, Côte d’Ivoire. Disponible sur: http://www.abidjan.district.ci/

37. Dongo K, Kouamé FK, Koné B, Biém J, Tanner M, Cissé G. Analyse de la situation de l’environnement sanitaire des quartiers défavorisés dans le tissu urbain de Yopougon a Abidjan, Côte d’Ivoire. VertigO. 8 janv 2009;(Volume 8 Numéro 3). doi:10.4000/vertigo.6252

38. Graham BL, Steenbruggen I, Miller MR, Barjaktarevic IZ, Cooper BG, Hall GL, et al. Standardization of Spirometry 2019 Update. An Official American Thoracic Society and European Respiratory Society Technical Statement. Am J Respir Crit Care Med. 15 oct 2019;200(8):e70–88. doi:10.1164/rccm.201908-1590ST

39. Quanjer PH, Stanojevic S, Cole TJ, Baur X, Hall GL, Culver BH, et al. Multi-ethnic reference values for spirometry for the 3-95-yr age range: the global lung function 2012 equations. Eur Respir J. déc 2012;40(6):1324–43. doi:10.1183/09031936.00080312 PubMed PMID: 22743675; PubMed Central PMCID: PMC3786581.

40. Boulet LP, Milot J, Boutet M, St Georges F, Laviolette M. Airway inflammation in nonasthmatic subjects with chronic cough. Am J Respir Crit Care Med. févr 1994;149(2):482–9. doi:10.1164/ajrccm.149.2.8306050

41. Adeloye D, Chan KY, Rudan I, Campbell H. An estimate of asthma prevalence in Africa: a systematic analysis. Croat Med J. déc 2013;54(6):519–31. doi:10.3325/cmj.2013.54.519 PubMed PMID: 24382846; PubMed Central PMCID: PMC3893990.

42. Tabka Z, Aouichaoui C, Debbabi F, Ben Dhiab W, Trabelsi Y, Abar I. Prévalence de l’asthme et des symptômes évocateurs d’asthme chez l’enfant et l’adolescent Djiboutien. Rev Fr Allergol. 1 juin 2020;15ème Congrès Francophone d’Allergologie 60(4):359. doi:10.1016/j.reval.2020.02.163

43. Jestin-Guyon N, Ouaalaya H, Tiaray Harison M. Impact of biomass fuel smoke on respiratory health of children under 15 years old in Madagascar. Respir Med Res. 1 juin 2023;83:100989. doi:10.1016/j.resmer.2023.100989

44. Kouao R, Kouame K, Silue s. Prevalence of asthma in children under 5 years old exposed to air pollution in Abidjan, (Côte d’Ivoire). ResearchGate. 2019. doi:10.24327/ijrsr.2019.1007.3524

45. Refiloe M, Kevin M, Rebecca N, Maia L, Hellen M, Graham D, et al. Asthma care in sub-Saharan Africa: Mind the gap! J Pan Afr Thorac Soc. 2 mai 2022;3(2):59–62. doi:10.25259/JPATS_12_2022

46. Magwenzi P, Rusakaniko S, Sibanda EN, Gumbo FZ. Challenges in the diagnosis of asthma in children, what are the solutions? A scoping review of 3 countries in sub Saharan Africa. Respir Res. 19 sept 2022;23(1):254. doi:10.1186/s12931-022-02170-y

47. Onisor MO, Turner S. Routine FEV1 measurement is essential in diagnosis and monitoring of childhood asthma: myth or maxim? Breathe. 15 août 2023;19(2):230048. doi:10.1183/20734735.0048-2023 PubMed PMID: 37645020.

48. Caillaud D, Annesi-Maesano I, Bourin A. La pollution atmosphérique et ses effets sur la santé respiratoire en France. Document d’experts du Groupe Pathologies pulmonaires professionnelles environnementales et iatrogéniques (PAPPEI) de la Société de pneumologie de langue française (SPLF). Rev Mal Respir. 1 déc 2019;36(10):1150–83. doi:10.1016/j.rmr.2019.10.004

49. Zhang W, Ma R, Wang Y, Jiang N, Zhang Y, Li T. The relationship between particulate matter and lung function of children: A systematic review and meta-analysis. Environ Pollut. 15 sept 2022;309:119735. doi:10.1016/j.envpol.2022.119735

50. Ngamsang P, Wongta A, Kawichai S, Kosashunhanan N, Chuljerm H, Khiaolaongam W, et al. Restrictive Lung Function Patterns and Sex Differences in Primary School Children Exposed to PM2.5 in Chiang Mai, Northern Thailand. Int J Environ Res Public Health. oct 2025;22(10):1530. doi:10.3390/ijerph22101530

51. Martinez FD. Early-Life Origins of Chronic Obstructive Pulmonary Disease. Drazen JM, éditeur. N Engl J Med. sept 2016;375(9):871–8. doi:10.1056/NEJMra1603287

52. Agustí A, Hogg JC. Update on the Pathogenesis of Chronic Obstructive Pulmonary Disease. N Engl J Med. 26 sept 2019;381(13):1248–56. doi:10.1056/NEJMra1900475

53. Hasan Md, Tasfina S, Haque SMR, Saif-Ur-Rahman KM, Khalequzzaman Md, Bari W, et al. Association of biomass fuel smoke with respiratory symptoms among children under 5 years of age in urban areas: results from Bangladesh Urban Health Survey, 2013. Environ Health Prev Med. 2019;24:65. doi:10.1186/s12199-019-0827-3 PubMed PMID: 31775610; PubMed Central PMCID: PMC6882069.

54. Hussein H, Shamsipour M, Yunesian M, Hasanvand MS, Mahamudu T, Fotouhi A. Fuel type use and risk of respiratory symptoms: A cohort study of infants in the Northern region of Ghana. Sci Total Environ. 10 févr 2021;755:142501. doi:10.1016/j.scitotenv.2020.142501

55. Adeyeye SAO, Oyewole OB. An Overview of Traditional Fish Smoking In Africa. J Culin Sci Technol [Internet]. 2 juill 2016 [cité 30 sept 2025]. Located at: world. Disponible sur: https://www.tandfonline.com/doi/abs/10.1080/15428052.2015.1102785

56. Pajot A, Yapo M, Liousse C. Feasibility and results of joint ambulatory monitoring of exposure to particulate matter pollution and lung function in children in Abidjan, Côte d’Ivoire, a cross-sectional observational study | medRxiv [Internet]. 2025. 10.1101/2025.08.16.25333819

57. Oluwole O, Arinola GO, Huo D, Olopade CO. Household biomass fuel use, asthma symptoms severity, and asthma underdiagnosis in rural schoolchildren in Nigeria: a cross-sectional observational study. BMC Pulm Med. 5 janv 2017;17(1):3. doi:10.1186/s12890-016-0352-8

58. Shin HH, Maquiling A, Thomson EM, Park IW, Stieb DM, Dehghani P. Sex-difference in air pollution-related acute circulatory and respiratory mortality and hospitalization. Sci Total Environ. 1 févr 2022;806(Pt 3):150515. doi:10.1016/j.scitotenv.2021.150515 PubMed PMID: 34627116.

59. WHO. WHO guidelines for indoor air quality: household fuel combustion [Internet]. Geneva: World Health Organization; 2014 [cité 15 avr 2022]. Disponible sur: https://apps.who.int/iris/handle/10665/141496

60. Bahino J, Giordano M, Beekmann M, Yoboué V, Ochou A, Galy-Lacaux C, et al. Temporal variability and regional influences of PM2.5 in the West African cities of Abidjan (Côte d’Ivoire) and Accra (Ghana). Environ Sci Atmospheres. 18 avr 2024;4(4):468–87. doi:10.1039/D4EA00012A

